# Understanding the impact of local authority resource allocation on gastrointestinal infections in England

**DOI:** 10.64898/2025.12.19.25342456

**Authors:** Lauren Murrell, Helen. E Clough, Xingna Zhang, Marie Anne Chattaway, Mark. A Green, Iain Buchan, Benjamin Barr, Daniel Hungerford

## Abstract

**Background:** Gastrointestinal infections are a substantial public health issue in England. Local authority environmental and regulatory (ER) services support the prevention and control of gastrointestinal infections with food safety and infection control functions. However, there have been significant and inequitable cuts to local authority budgets, with ER services seeing expenditure reduced by 2.4% per capita in the most deprived compared to 1.2% per capita in the least deprived authorities. It is therefore imperative to understand the impact local funding cuts to ER services may have on gastrointestinal infection outcomes.

**Methods:** We use longitudinal data in England, at local authority district level, between 2010 and 2019. Exposures of interest were ER spending lines of food safety expenditure, and aggregated spending lines of food safety and infection control (FSIC) expenditure, and the number of food hygiene full time equivalent staff (FTE) per 10,000 of the population. Primary outcomes of interest were the number of laboratory-confirmed Campylobacter, Salmonella, and E. coli O157 infections, emergency hospitalisations due to GI infection, and the number of calls to NHS 111, all at the local authority level. We use fixed effects negative binomial modelling to estimate the association between relative change in the incidence rate of gastrointestinal infection outcomes and increases in each of our exposures. NHS 111 and hospital admission data were desegregated by age group, and age-exposure interactions were used to understand differential effects. This was not possible for pathogen data due to the small number of observations per year by species.

**Results:** No significant relationship was found between expenditure or staff and foodborne pathogen incidence overall. When considered as a whole, we find no significant relationship between expenditure or staff levels and the number of hospital admissions for all age groups. However, when disaggregated by age, an increase of £1 per capita in food safety expenditure was associated with a 1.34 % decrease in the rate of hospitalisations among 20–59-year-olds (IRR = 0.9866: 0.9738, 0.9996). In addition, an increase in food hygiene staffing per 10,000 of the population was associated with a 36% decrease in the rate of hospitalisations in 60–64-year-olds (IRR =0.64: 0.48 0.86). Results also identified an increase in resource allocation was associated with increased rate in hospital admissions for some age groups. A £1 per capita increase in food safety expenditure was associated with a 3% increase in hospitalisations for 5-9 (IRR=1.0300: 1.0001, 1.0607) and a 2.3 % increase for 10-19-year-olds (IRR=1.023:1.004,1.043). £1 per capita increase in FSIC expenditure was associated with a 1% increase in admissions for 75+ (IRR=1.009:1.001,1.018). Finally, a one unit increase in staff per 10,000 of the population was associated with a 134% increase admission for 5-9-year-olds (IRR =2.34:1.60, 3.43) and 50% increase amongst 10-19-year-olds (IRR=1.50:1.14, 1.98), whilst no significant relationship was identified between the number of NHS 111 calls and exposures of interest.

**Discussion:** Overall, no significant relationship was identified between expenditure or staff levels on the number of laboratory confirmed pathogens, hospital admissions or NHS 111 calls. However, age-specific patterns suggest an increased resource allocation may be protective for some groups in preventing hospital admissions. While increased rates of illness for children and 75+ years may reflect differences in exposure, detection or healthcare seeking behaviour or other unknown factors. Building on previous research evidencing the inequal reductions in service expenditure, and the impacts on service capacity, these results highlight the potential role of local funding cuts on GI infection health outcomes. Further research is warranted to understand the mechanisms driving different effects and to understand how service provision interacts with sociodemographic factors.

## Introduction

In the UK, Gastrointestinal (GI) infections are common, with an estimated 17 million cases annually (1). They place a substantial burden on the National Health Service (NHS) with an estimated 1 million GP consultations a year, and 118,000 emergency hospital admissions in 2019-20 (1,2). Pathogens that cause GI infection are commonly spread person-to-person, by food and water, or contaminated surfaces or animals (3).

In England, the UKHSA (United Kingdom Health Security Agency) is the lead organisation responsible for the surveillance of GI infection pathogens, working with public health agencies and local authority colleagues (4,5). The UKHSA receive notifications for diseases such as *Campylobacter, Salmonella* and Shiga toxin producing *E.coli* O157 (STEC O157) (6). These are three out of four key pathogens that are part of routine surveillance due to large numbers of cases (*Campylobacter* and *Salmonella)* or high severity of illness (*STEC O*157) (7). In addition, these pathogens are the most common bacteria that contaminate food, along with *listeria monocytogenes* (4). *Campylobacter* is the most commonly identified source of acute diarrhoea in the UK, with an estimated 300,000 cases acquired by food out of approximately 630,000 total cases in the UK annually (8,9). *Salmonella* can be typhoidal or non-typhoidal, typhoidal is often associated with travel to endemic countries (10). Non-typhoidal *Salmonella* are often associated with foodborne outbreaks, and are primary cause of foodborne disease, causing 27% of all foodborne outbreaks between 2015 and 2020 (11). Illness with *E.coli* O157 can causing severe disease and is found in foods such as under-cooked meats, raw milk and vegetables (12). The Food Standards Agency (FSA) estimates that 2.4 million cases of foodborne illness occur each year (13). An estimated 180 deaths a year are attributed to 11 key foodborne pathogens (14). Norovirus is responsible for the most deaths with an estimated 56 deaths a year, followed by *Salmonella* causing approximately 33 deaths a year in the UK (14).

Local authorities provide services that aim to protect public health. Environmental and regulatory (ER) services in England are responsible for providing services such as trading standards, waste management, water safety, food safety and animal public health and infectious disease control (15). In England the FSA is responsible for food control and works with local authorities which are responsible for the enforcement of food hygiene regulations (16). The responsibilities of local authorities and their food safety teams include advising consumers and businesses on issues pertaining to food, inspections for food hygiene or standards, sampling, and advice visits, to check and ensure compliance with food law (16,17). Food safety activities to reduce food and water-based poisoning, fall under the food safety spending line (15). The animal and public health and infectious disease control spending line includes functions of public health protection such as cesspool emptying, sewerage scheme work, health education and infectious disease control under the Public Health (Control of Diseases) Act of 1984, and associated regulations (15). Examples of functions that may be conferred on local authorities include the monitoring of public health risks and imposing restriction in relation to a public health threat (18).

Since the introduction of cuts in 2010, local authorities have seen reductions to their funding by up to 50% (19). Previous research showed that ER services have faced reductions in expenditure which have been largest in the most deprived areas, experiencing reductions in expenditure per capita of 2.4% compared to 1.2% in the least deprived areas (20). Research also showed that food safety and infection control (FSIC) services saw local funding cuts of up to 23% in the most deprived areas compared to 8% in the least deprived areas (21). In addition, a recent study carried out on data between 2009/10 and 2019/20, demonstrated that reductions in food safety expenditure were significantly associated with decrease in food hygiene staff numbers, and the number of interventions carried out per establishment (22). It is unclear what effect these cuts may have on food safety-related public health. In recent years there have been reports of increased emergency hospitalisations due to foodborne disease (23). However, there is no research to our knowledge that focuses on the impact of local government expenditure reductions, and GI infection outcomes. This study aimed to understand the impact of resource allocation to local authority services on gastrointestinal infection outcomes.

## Methods

We conducted a longitudinal ecological study at the lower tier local authority level using annual data from 2009 to 2019. The study was based on 314 local authorities in England, using the 2020 geographic boundaries. The Isles of Scilly and the City of London were removed due to distinct characteristics of small population sizes and distinct funding structure. A published protocol contains more detail on the data (24), and we cover more detail on local authority structures in previous study (21). Thirteen local authorities were removed due to missing food safety or food safety and infection control expenditure. A further five were excluded due to outturns reported with other functions outside of food hygiene. Six local authorities were excluded due to inconsistent reporting. In total 26 were removed due to data quality issues leaving 288 carried forward for analysis. More detail can be found in the supplementary material section 2.

### Data

#### Gastrointestinal infection indicator data

We use three primary outcome indicators of gastrointestinal infections from three data sources.

First, we used the number of positive laboratory-confirmed cases of key gastrointestinal infection pathogens which are often associated with foodborne transmission. These pathogens were *Campylobacter, E. coli* O157 *and Salmonella.* We excluded *Salmonella* Typhi and Para typhi from the analysis as they are largely associated with travel and our focus is on home-acquired infection. The data were analysed at the local authority level, as the number of confirmed cases each year. Data were provided by UKHSA, previously Public Health England. Laboratory confirmed pathogen detections are held on central data base within UKHSA called the Second-Generation Surveillance System (SGSS) (6). More Information on how data are reported can be found in the guide for diagnostic laboratories (6). Data were not analysed by age due to statistical disclosure concerns regarding small observation counts for each pathogen when broken down by age and year.

Secondly, the Hospital Episode Statistic data (HES) from NHS Digital, provides data on the number of emergency hospital admissions to NHS hospitals due to infectious intestinal disease (25). Admissions are defined using ICD-10 Codes A00*-A09* + K52.9, excluding A09.9 and K52.9 which represent unspecified or non-infectious gastroenteritis and colitis. We group the data to local authority level and create age groups of 0-5, 5-9, 10-19, 20-59, 60-74 and 75+. Data were analysed as the number of emergency hospital admissions each year. More information on this data can be found on the HES website (25). We use this measure to examine more severe outcomes of illness in the community.

Third, we used syndromic call data for diarrhoea and vomiting collected from the NHS 111 telephone helplines. This provides information on community incidence, reporting calls for symptoms of GI infection (diarrhoea and vomiting as the primary presenting symptoms) (26). NHS 111 helpline, a successor of NHS Direct, was established in 2013, and provides guidance over the phone, offering an alternative form of point of care. NHS 111 data are reported at the postcode district level or upper tier local authority level, of which there are 152 authorities. Our analysis was at lower tier level. For this reason, we were only able to successfully link upper tier data to lower tier local authority level in cases where local authorities were single tier, non - county authorities. We were able to do this for 124 authorities. The data were provided in age groups of 0-4, 5-9,10-15, 16-19, 20-59, 60-74, 75 + years. For consistency and ability to match age groups with HES data, age groups 10-15, and 16-19 were grouped to form a single group 10-19 years. The number calls for vomiting and diarrhoea were combined to form one variable.

For HES and NHS111 data sources, we removed data for 0–4-year-olds from analyses. In this age group rotavirus was the predominant cause of GI illness until rotavirus vaccination was introduced for infants in 2013 (27,28). The introduction of the vaccine drastically decreased GI illness in this age group (27). Therefore, to account for this confounding effect this age group is removed from all analyses.

Collectively, these data sources offer insight to the incidence of GI infections and enable monitoring and surveillance of trends within different sectors and at varying degrees of disease severity.

### Expenditure data

Our primary exposures of interest were expenditure of spending line of food safety and the aggregated spending line of food safety and animal public health and infection control, which we will refer to as infection control (IC), to form food safety and infection control (FSIC). This aggregated line allowed us to capture if functions related to spending on infection control under this spending line may be associated with GI infection outcomes in combination with the food safety activities. Furthermore, due to potential misreporting, discussed in previous work (22), this approach increases the ability to capture spending related to prevention of GI infection. The expenditure data were sourced from the Place-based Longitudinal Data Resource (PLDR) (29). This data resource collates publicly available expenditure data on local authority annual outturn for cultural, environmental and regulatory services, reported to the UK central government department of housing and local government (30). Expenditure data were adjusted for inflation using gross domestic product deflator (31), then normalized by ONS mid-year population estimates to give food safety services and FSIC expenditure *per capita*.

### Local authority service indicator data

Our secondary exposure of interest was the number of full-time equivalent (FTE) food hygiene staff. Data were obtained from the FSA’s local authority enforcement monitoring system (LAEMS) (32). We used the ONS mid-year population estimates to derive the number of FTE per 10,000 of the population as the exposure variable used in analysis. This measure allows us to understand how food hygiene staff and their activities relate to gastrointestinal infection outcomes, allowing for population density. More information on this measure and its limitations can be found in supplementary material section 1.

### Population data

Local authority level population estimates were obtained from the Office for National Statistics (ONS). Deprivation data was obtained from English Indices of Multiple Deprivation (IMD) from the Ministry of Housing, Communities and Local government (33). The IMD was included in the model to account for area-level socioeconomic deprivation.

### Missing data

There were missing data present in the exposure variables of interest, namely food safety and infection control expenditure, FTE, and the number of interventions achieved. Local authorities missing over 50% of data for any indicator were removed. In some cases, local authorities reported food standard alongside food hygiene or only food standards returns – these authorities were removed. In addition, local authorities with inconsistent reporting were removed. The remaining 288 local authorities were carried forward for multiple imputation using the R MICE package (https://cran.r-project.org/web/packages/mice). We carried out sensitivity analysis on data prior to multiple imputations. More detail can be found in the supplementary material (S3.1).

### Analysis

First, we descriptively analysed the data to visualise annual trends in the outcome indicators of interest by socio-economic characteristics and age.

We then used fixed effects negative binomial regression to model the association between change in our exposures and change in our outcome variables. The incidence rate ratio (IRR) was obtained by exponentiating the model coefficients to interpret the multiplicative change in outcome per unit increase in exposure. The negative binomial family was used to model the count data used and to accommodate overdispersion detected in the data. Local authority fixed effects were included to remove all between local authority differences to allow models to estimate the associations present between exposure and outcomes within local authorities over time. All models also used robust standard errors to account for clustering within local authorities. In addition, all models included deprivation as an interaction term with year, which allowed us to account for differences in temporal trends by deprivation over time. For each model, the log of the population size was included as an offset in order to model the rate and the model was weighted for population to adjust for differences in population across areas overtime.

As NHS 111 and hospital admission outcomes were disaggregated by age we additionally included age-exposure interactions to allow for the model to account for baseline differences in calls by age and how the effect of the exposure varies by age group. When modelling hospital admissions, we also adjusted for rotavirus vaccine introduction in 2013, including a binary variable 0 for before introduction and uptake and 1 as post 2013 introduction and uptake. As the NHS111 data was only available post 2013, this binary variable was not included in these models.

While the narrative of this paper focuses on the effect of reductions in expenditure, we report the results in terms of unit increase in exposure. This perspective allows us to estimate the marginal effect of re-investment of expenditure and staff in relation to GI infection health outcomes. By framing our results this way, we highlight the value of resource allocation which in turn facilitates understanding of the potential benefits of re-investment or alternatively the prevention of further cuts. This allows our estimates to reflect the positive outcomes of investment in the context of local funding cuts. All results are presented as main effect and 95% confidence interval unless otherwise stated.

### Supplementary analysis

In addition to sensitivity analysis (section S3.1) supplementary analysis includes NHS111 data for months June to December, excluding peak norovirus months (Section S3.2). Section S3.3 compares outcomes in the least deprived authorities (Q1) and the most deprived (Q5). Section 3.4 includes analysis using ER expenditure and the total number of interventions achieved per establishment as exposures of interest. In addition, we carried out analysis including a 1-year lag on exposures of interest, to see the effect of previous year spending and staffing on outcomes of interest, more can be found in S3.5.

## Results

Table 1 describes the change in exposure variables of interest between the beginning and end of the study period. The average food safety expenditure decreased from £3.14 to £2.27 between 2009 and 2019. The 2009 average FSIC expenditure was £5.48 higher than in 2019. The average number of FTE per 10,000 decreased from 0.31 to 0.24.

**Table 1:** Descriptive summary table of exposure variables of interest.

| Year | Average FS expenditure per capita | Average FSIC expenditure per capita | Average FTE per 10, 000 of the population |
| --- | --- | --- | --- |
| **2009** | £3.14 | £9.05 | 0.31 |
| **2010** | £3.20 | £8.08 | 0.30 |
| **2011** | £2.85 | £6.57 | 0.29 |
| **2012** | £2.79 | £5.88 | 0.28 |
| **2013** | £2.66 | £5.11 | 0.27 |
| **2014** | £2.45 | £4.70 | 0.27 |
| **2015** | £2.44 | £4.55 | 0.26 |
| **2016** | £2.34 | £4.16 | 0.25 |
| **2017** | £2.32 | £3.98 | 0.25 |
| **2018** | £2.30 | £3.74 | 0.25 |
| **2019** | £2.27 | £3.57 | 0.24 |

Figure 1 shows the rate of laboratory confirmed pathogens between 2009 and 2019 per 100,000 of the population. All pathogens experienced a decline in laboratory confirmation rate between 2009 and 2019, with some fluctuation over time. *Campylobacter* had the largest incidence during this time, increasing from 113 to 116 per 100,000 of the population in 2012 before returning to just below the baseline rate in 2019. *Salmonella* experienced an overall decline in laboratory confirmed cases, decreasing from 17 to 14 per 100,000 in 2019 with some fluctuation over time. *E. coli O157* occurred at much smaller rates, peaking at 2.4 per 100,000 in 2011 before declining to 0.8 per 100,000 by 2019.

**Figure 1.**
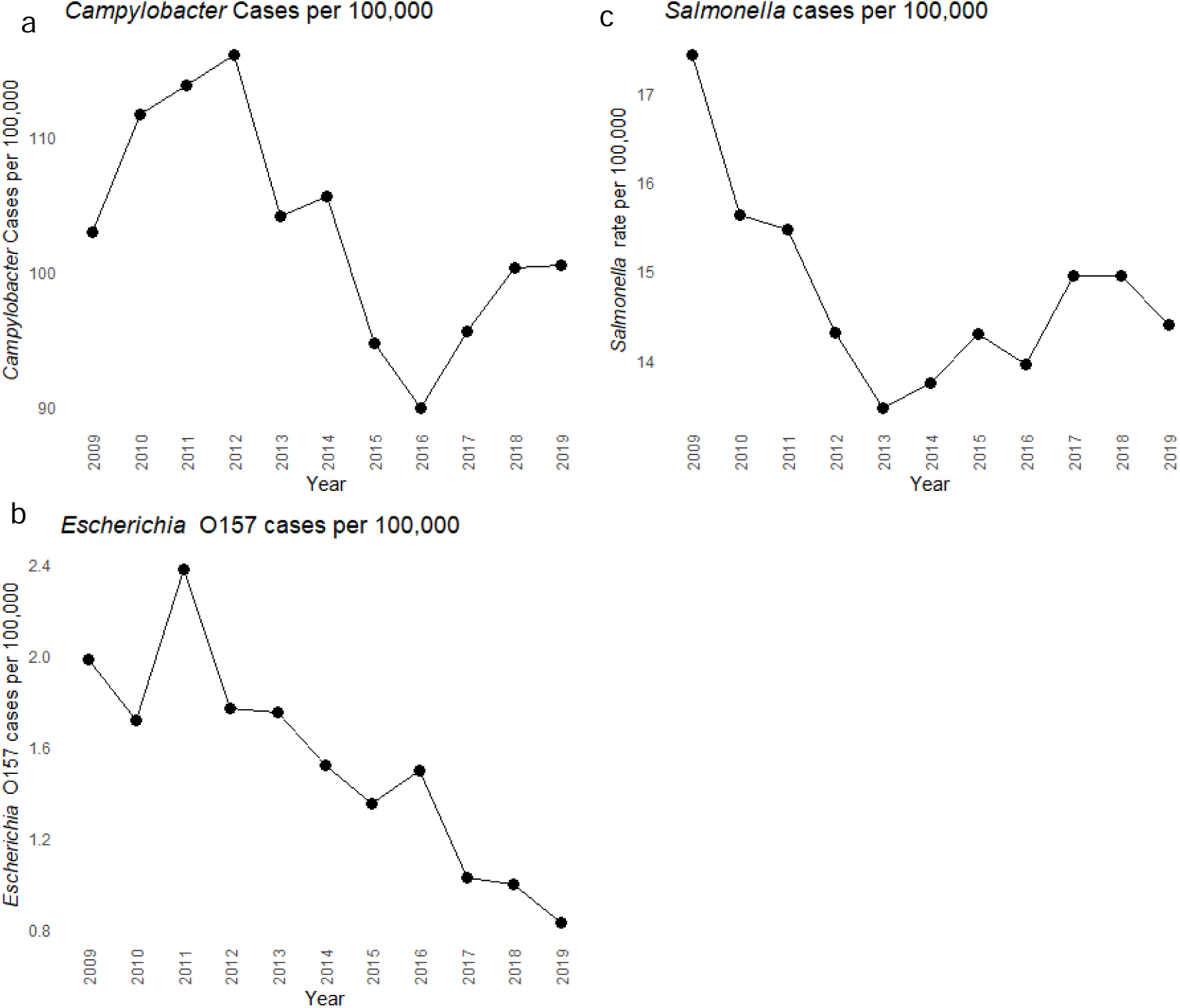
Rates of laboratory confirmed Campylobacter spp. (figure 1a), E. coli O157. (figure 1b) and Salmonella spp. (figure 1c) (excluding typhi and para typhi) per 100,00 of the population of 2009-2019.

Figure 2 shows the rate of hospital admissions for GI infection per 100,000 stratified by age group between 2009 and 2019. Admissions among 0-4-year-olds declined substantially from 773 to 470 per 100,000. All other age groups experience an increase in the rate of hospitalisation during this time. Those aged 5-9-years-old experience the largest increase from 102 to 152 per 100,000. Other age groups experienced relatively smaller increases in admission rate, with those aged 20-59 experiencing the lowest rate of hospital admission during this period.

**Figure 2.**
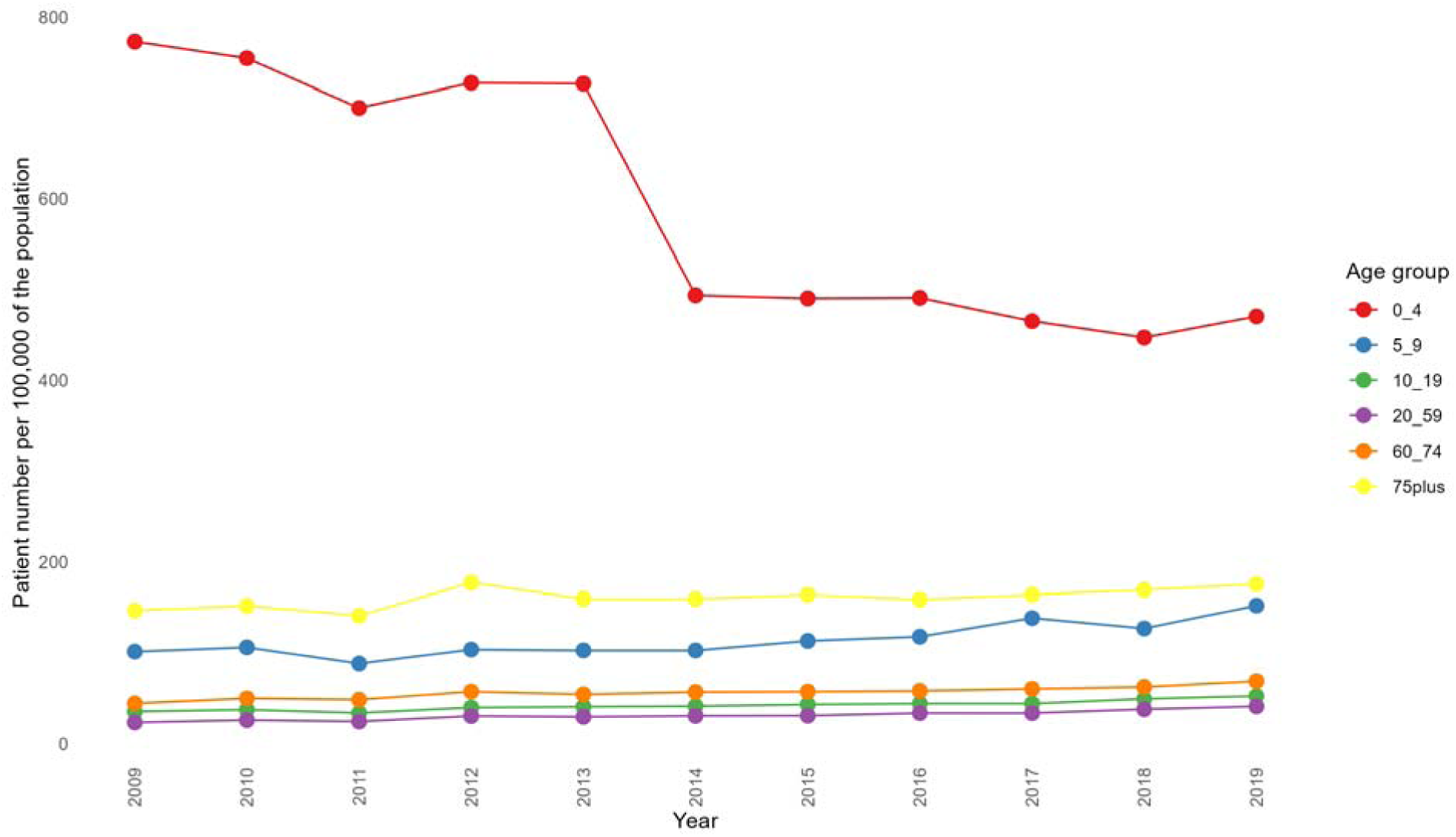
The rate of hospital admissions for GI disease per 100,000 of the population by age group 2009-2019.

Figure 3 shows the number of calls for vomiting and diarrhoea between 2014 and 2019. The highest rate of calls was for 0-4-year-olds, increasing from 3476 to 4162 per 100,000 between 2014 and 2016, before dropping to 4010 per 100,000 in 2019. This was followed by the 75+ group which experienced little variation over time, increasing from 735 to 772 per 100,000. The 5-9-year-old group experienced an increase in calls from 465 to 657 per 100,000. The other age groups: 10-19-year-olds, 20-59-year-olds, 60-64-year-olds and 75+ year-old cohorts experience relatively smaller increases between 2014 and 2019.

**Figure 3.**
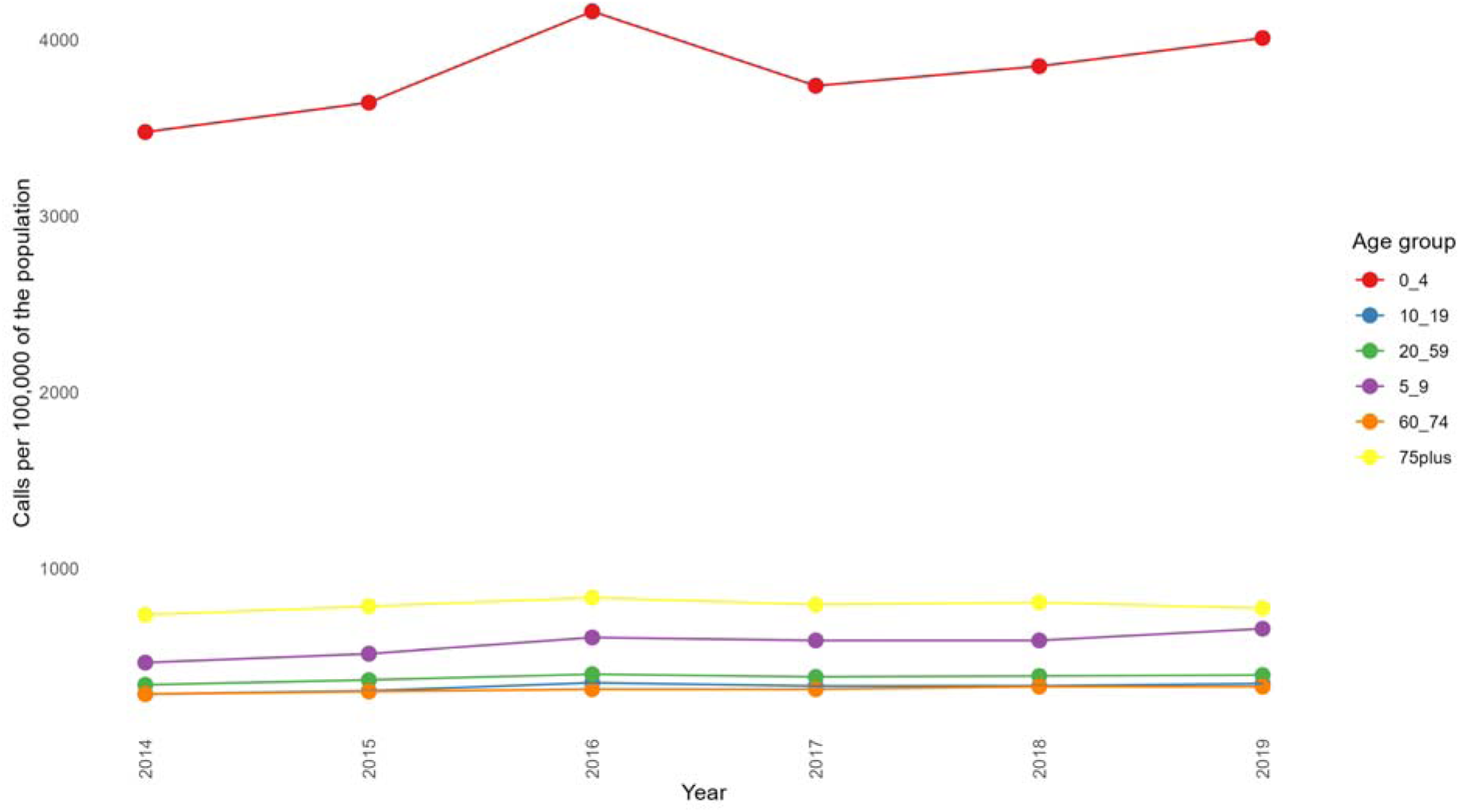
Number of calls to NHS 111 for diarrhoea and vomiting by age group per 100,000 of the population 2014 – 2019.

### Statistical analyses

The Incidence Rate Ratio (IRR) of laboratory-confirmed foodborne pathogens estimated using negative binomial fixed effects regression are reported in Table 2. There was no significant association between rates of laboratory-confirmed *Campylobacter, Salmonella,* or *E. coli* O157 and food safety expenditure, FSIC expenditure, or FTE.

**Table 2:**
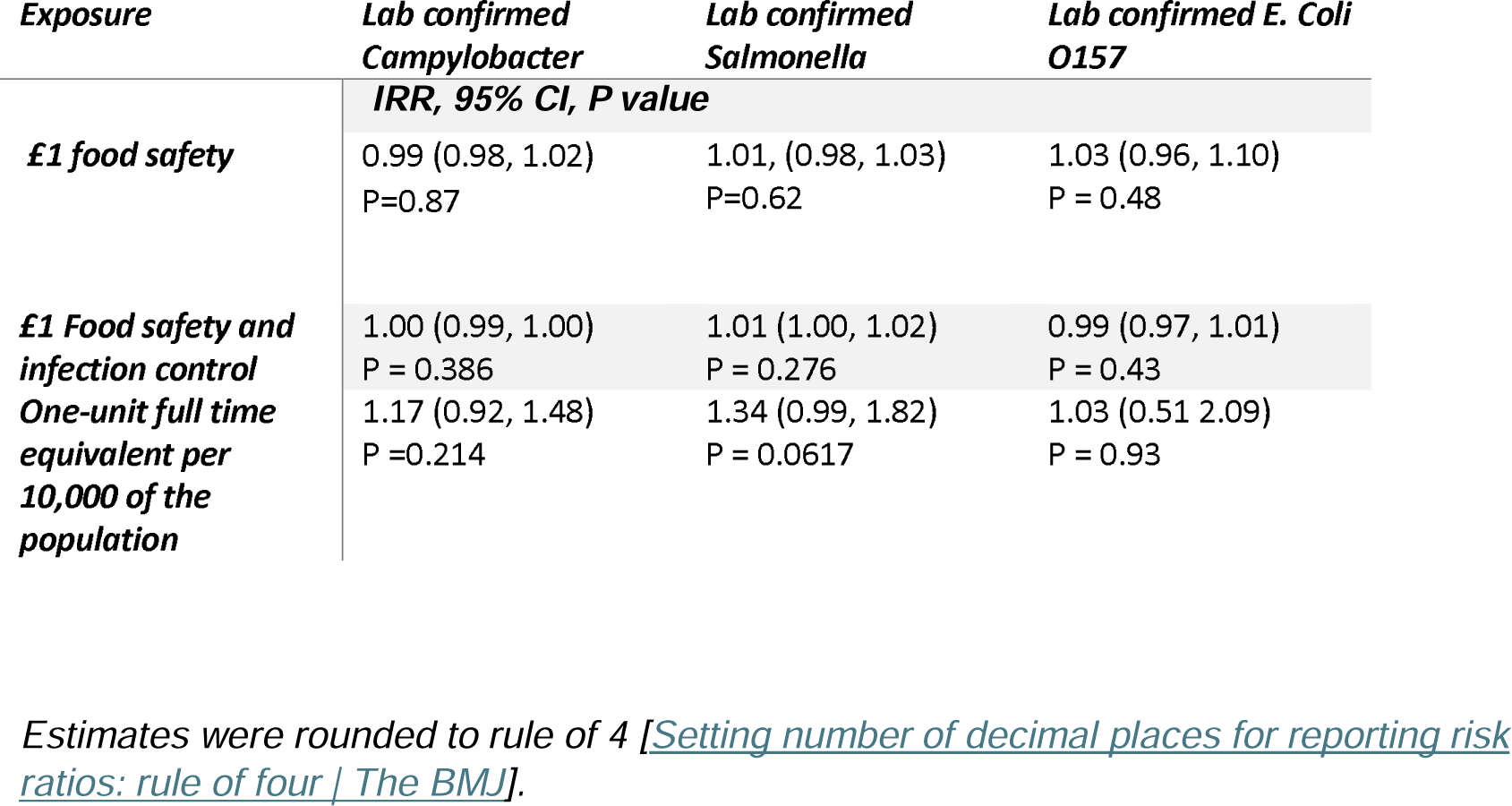
Incidence rate ratio (95% confidence interval) and P values for laboratory confirmed pathogens by species-based associated with service expenditure and staff (FTE).

Deprivation-stratified analysis reported in supplemental material section S3.3 identified that every £1 increase in food safety expenditure was associated with a reduced rate of laboratory-confirmed *Campylobacter* in the most deprived areas (IRR=0.96: 0.94, 0.99) and a decrease in *Salmonella* in the least deprived areas (IRR=0.95: 0.91, 0.99). A £1 increase in FSIC expenditure was associated with a decrease in *E. coli* O157 (IRR=0.968: 0.937, 0.999) in the least deprived areas. One unit increase in staff was associated with a decrease in *E. coli* O157 (IRR=0.092: 0.0171, 0.49) in the least deprived areas (see supplementary material section 3.3 for more detail.)

Table 3 shows the IRR of hospital admission estimate from fixed effects negative binomial model.

**Table 3:**
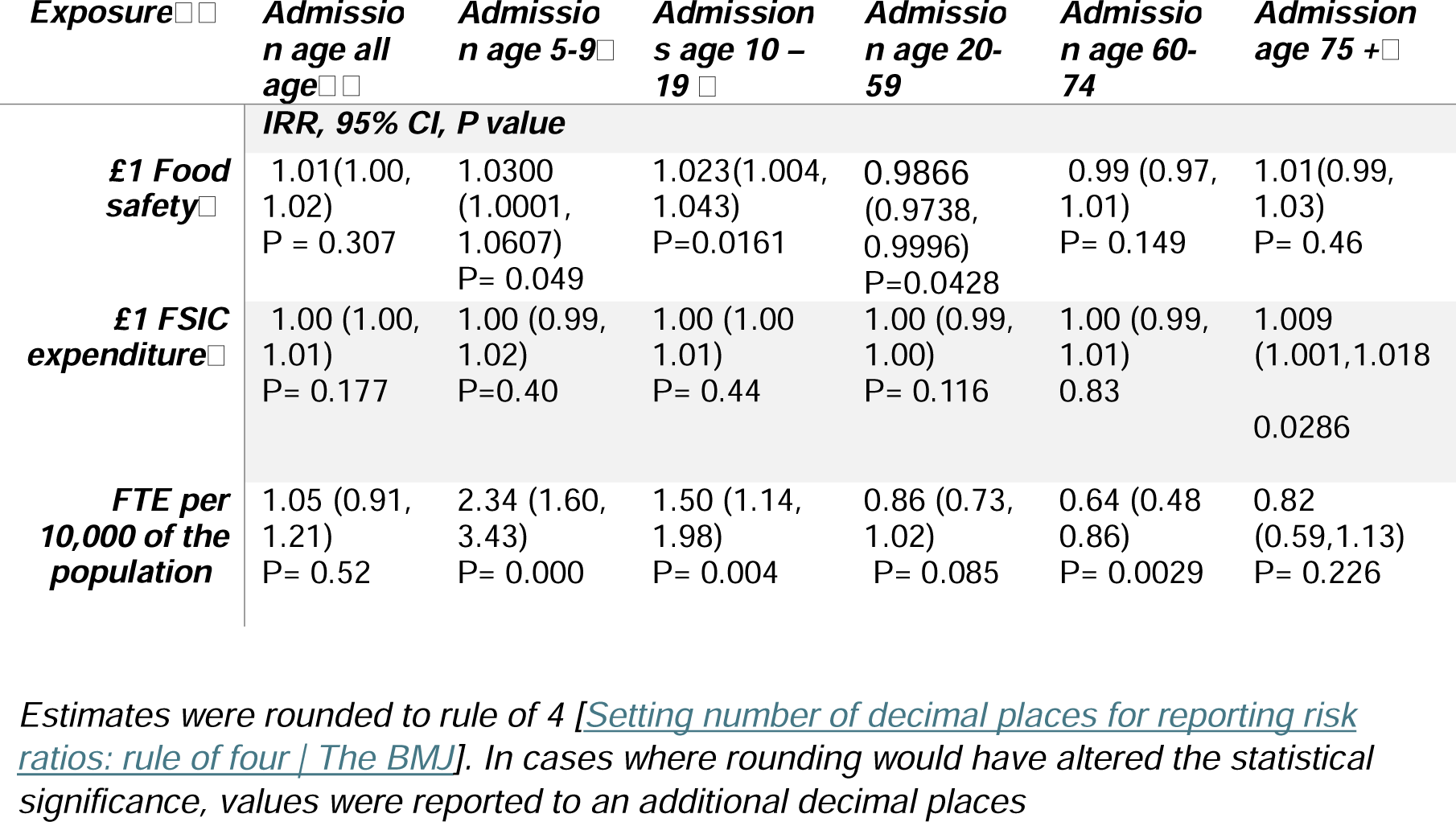
Incidence rate ratio (95% confidence interval) and P values for hospital admission by age group, associated with service expenditure and staff (FTE).

A £1 increase in food safety was associated with a lower rate of hospital admissions among 20–59-year-olds (IRR = 0.9866: 0.9738, 0.9996). In contrast, a £1 increase in food safety expenditure was associated with a higher in rate of admissions among 5-9-year-olds (IRR=1.0300: 1.0001, 1.0607) and among 10-19-year-olds (IRR=1.023:1.004,1.043). A £1 increase in FSIC expenditure was significantly associated with an increased rate of hospital admissions among those aged 75+ years (IRR=1.009:1.001,1.018). A one unit increase in FTE per 10,000 of the population was associated with a lower rate of hospital admissions among 60–74-year-olds (IRR =0.64: 0.48 0.86). Conversely, one-unit increase in staff was associated with a higher rate of hospital admissions for those aged 5-9 (IRR =2.34:1.60, 3.43) and 10-19-year-olds (IRR=1.50:1.14, 1.98). No significant relationships were identified between food safety, FSIC expenditure, or food hygiene staff and the rate of hospital admissions, or calls when stratified by deprivation, see supplementary material section S3.3.

Tabel 4 reports the IRR based upon NHS 111 data which was estimated using fixed effects negative binomial modelling. We found no association between food safety, FSIC or staff and the rate of NHS 111 calls. In addition, no significant relationships were identified between food safety, FSIC, or food hygiene staff and the rate of NHS 111 calls when stratified by deprivation (supplementary material 3.3).

**Table 4:**
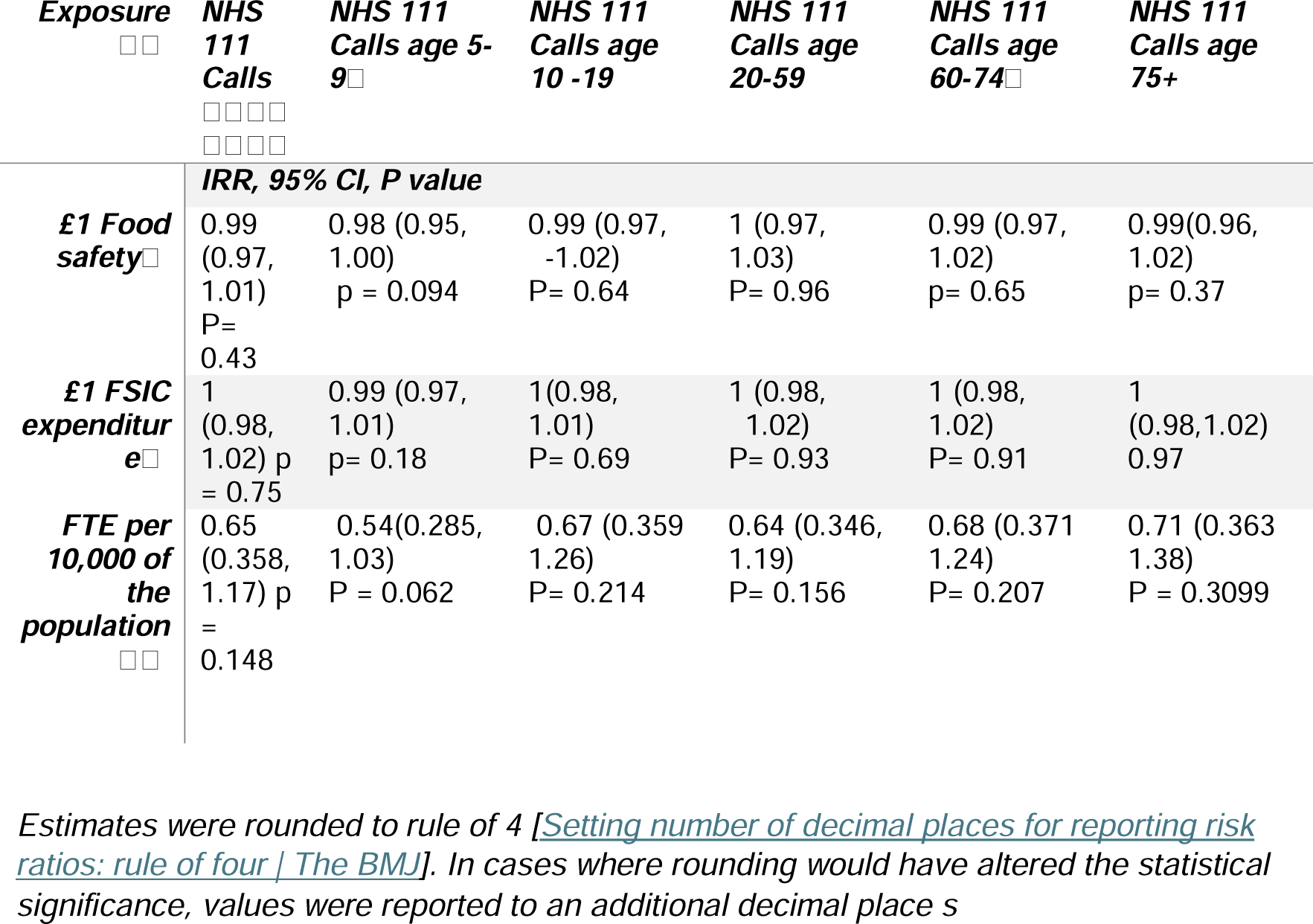
Incidence rate ratio (95% confidence interval) and P values for NHS 111 call by age group, associated with service expenditure and staff (FTE).

## Discussion

### Summary

Our results show no clear or consistent pattern to the relationship between local authority resource allocation and GI infection outcomes. The results reveal a complex association between exposures and outcome when analysed by age groups. Increased expenditure and staffing were associated with age-dependent effects for hospital admissions for GI infections. Specifically, an increase in food safety expenditure was associated with a reduced rate in hospital admissions amongst the 20-59-year-olds, while one unit increase in staff was associated with decreased hospital admissions in 60-74-year-olds. Conversely, increased food safety expenditure and staffing were associated with higher rates of hospitalisation in 5-9 and 10-19-year-olds. In addition, an increased FSIC expenditure was associated with an increased rate in hospital admissions in 75+.

When stratified by deprivation a £1 increase in food safety expenditure was associated with a significant reduction in the rate of *Campylobacter* cases in the most deprived areas. In contrast increased food safety expenditure in the least deprived areas was associated with decreased *Salmonella* cases, while increased FSIC expenditure and staffing were associated with a decrease in the rate of *E. coli* O157 case rate in the least deprived authorities.

### Interpretation

Laboratory-confirmed GI pathogens reflect only a fraction of the true incidence in the community. The reporting of GI infection for surveillance rests on individual’s healthcare seeking behaviours. Laboratory data do not reflect community level rates, but more likely those experiencing more severe illness (3,34). This underestimation of the true incidence may explain the lack of association identified between resource allocation to local authorities and laboratory reported rates.

Findings from deprivation-level analyses showed that increased food safety expenditure was associated with decreased laboratory-confirmed *Campylobacter* in the most deprived areas. This may indicate that resource allocation is more impactful for foodborne disease caused by *Campylobacter* in areas of higher deprivation. However, it is important to note the results may be biased due to reporting practices from different areas varying by the level of deprivation (35). Evidence suggests that individuals form more deprived areas with *Campylobacter* are less likely to be reported to surveillance (36), leading to under ascertainment in this cohort. In addition, the most deprived areas faced the largest reductions in funding, which may have influenced testing and therefore reporting, and may have contributed to underestimation of cases in more deprived areas. Therefore, bias may be introduced wherein the results show a drop in cases associated with increased food safety expenditure, inflated by under-reporting associated with local funding cuts. In contrast the rate of S*almonella* detections decreased with increase food safety expenditure in the least deprived areas and *E. coli* O157 showed a similar association with an increase in FSIC expenditure and staffing levels. The results suggest that resource allocation may have a more pronounced impact in the less deprived areas where local funding cuts are smaller (21) and there may be a greater capacity for surveillance, reporting and intervention.

Our results show that, increased food safety expenditure is associated with a decreased rate of hospitalisations in amongst 20-59-year-olds. In addition, one unit increase in staff is associated with a decreased rate of hospitalisations in 60-74-year-olds. These results offer evidence that indicates the importance of public health investment in the prevention of GI infection illness. The investment of resources in local public health teams has been shown to be an effective way to reduce foodborne illness. A US-based demonstrated that an increased capacity of local environmental health teams and food protection programs are protective against foodborne illness, with counties that had larger budgets experiencing reduced illness (37). In addition, workforce capacity has been associated with less illness, with counties with the highest FTE experiencing 36% lower rate in foodborne illness. This identifies some consistency with our findings in providing evidence of the role of staff in the protection of GI infection and illness.

However, increased resource allocation did not show a consistent protective association across all age groups, the reasons for this are less clear and may be partly explained by different factors. Differences in exposure to food safety setting may contribute to these findings. Adults aged 75 and over report frequenting pubs, restaurants and takeaways the least out of all age groups surveyed (38). This implies that increased expenditure and staffing may provide greater protection for those more frequently exposed to these settings (20-59- and 60-64-year-olds). Explanations behind trends seen in school aged children are less clear. Children are acknowledged to have poorer levels of hygiene, in addition to exposure to areas with potential for transmission or outbreak such as school (39,40). This may mean an increase food safety resource allocation may be less influential on health outcomes. Therefore, health outcomes in these age groups may remain high despite increased food safety resource allocation. In addition, differences in susceptibility to severe illness and hospitalisation may contribute to higher rates of these age groups reporting to or being referred to healthcare. Both younger children and older adults tend to have weaker immune systems making them more susceptible to infection and illness (41–43). Older adults face an increased vulnerability to GI illness with higher incidence of hospitalisation and length of stay due to GI illness (44). In addition, older age groups may report more to healthcare than other age groups, resulting in higher rates or reported illness (45,46). Another consideration is that the reductions in funding may lead EHO’s to prioritise more vulnerable groups, increasing ascertainment and referral rates within these cohorts.

It is also possible that our findings reflect reverse causality present, whereby areas with a higher rates of GI illness and hospitalisation drive the need for public health intervention requiring expenditure and staff allocation in response.

Overall, our findings may reflect a combination of factors, including variations in risk and exposure, behaviour, surveillance practices alongside the potential for reverse causality. In addition, there may be other explanations driving these results that we have not identified. The inconsistent results suggest that the relationship present between resource allocation and health outcome in GI infection is complex and requires further and more in-depth investigation before broad conclusions can be made.

Other factors influencing gastrointestinal infections were beyond the scope of this study. For example, viral pathogens mostly spread by person-to-person transmission such as Norovirus, which is highly infectious and is associated with outbreaks in schools, and nursing homes (47). Reductions in infection control expenditure may influence incidence rates of such pathogens, through the funding of activities related to outbreak control, sanitation, and advice. This represents an area for further study. In addition we do not account for illness due to sewerage, whilst the recent spillages reported in England have been identified as a growing public health problem (48), these events independent of local authority funding may influence our outcomes of interest and would be a useful line of further investigation.

### Strengths and limitations

A key strength of this paper is the use of different indicators of GI infection. This captures a wider range of outcomes at different levels of surveillance, therefore giving a better understanding of GI infection across the community. Further, we deliberately consider a range of outcome variables which represent a range of different severities of disease outcome, across a diverse geography and extended time window, and ours is the first study to provide such multi-perspective insights. It is also the first study to analyse trends in GI infection data in relation to local authority resource allocation, providing evidence that may help influence or inform policy at the local level. We show very effectively how routinely collected data which can be obtained at minimal cost can be used together with novel statistical methods to provide valuable insight into gastrointestinal disease risk within our population that can be used to inform health protection strategies.

There were several limitations. First, the NHS 111 data were only available for a subset of local authorities (single tier). While we attempted to estimate data for two-tier areas, we were no able to yield sufficiently reliable results. Consequently, any conclusions drawn from NHS 111 data excludes district authorities in two tier areas. In addition, NHS 111 started in England in 2014, reducing the years of data available for this indicator. In addition, there were missing data in exposure variables, to address this we used multiple imputation to estimate missing values.

Secondly, the laboratory data may not provide a representative picture of the rate of pathogens in the community and may underrepresent more socioeconomically deprived populations (49). In addition, we focus only on E. coli O157, other type of STEC are becoming an emerging threat and becoming a health burden, this could be focused on in future studies.

Third, NHS 111 and hospital data will include illness due to viral pathogens which largely transmits person-person. To mitigate this, we conducted sensitivity analysis excluding months where Norovirus peaks, which was only possible for NHS 111 data as hospital data was not provided by month. We also accounted for introduction of the rotavirus vaccine in 2013 by including a post-2013 binary variable hospital models (all NHS111 data were post 2013). In addition, removal of 0-4-year-olds from both data sets to help account for the impact of the rotavirus vaccine.

Finally, it is also possible that the outcomes measures are influenced by time varying unmeasured confounding factors, such as changes in public health policy which might influence both resource allocation and GI illness. Additionally, we use ecological observational study design, using area level data. Results may therefore be affected by ecological fallacy, in that trends at the aggregated local authority level may not reflect results at the individual level. Assumptions at this level may therefore introduce bias as inferences at the individual level cannot be made.

### Implications for policy and practice

Our work follows a period of sustained funding cuts in England, which have seen local authority budgets substantially reduced especially in deprived areas (19,21,50). Specifically cuts to ER and FSIC services are largest in the poorest authorities, single tiers and associated with increasing population density. During these cuts, food safety teams have reported reduction in capacity, staffing, staff experience, inspections and increased delays (51,52). Food safety expenditure reductions were significantly associated with staff reductions, and in turn reduction in the number of interventions achieved (22).

The capacity of local health departments has shown to be important in the timeliness and ability to investigate and report foodborne disease and outbreak (53,54). Consequently, the reduction of food safety, FSIC budgets and work force may place the public at greater risk of GI illness. This work provides evidence of the importance in resource allocation in the reduction of foodborne pathogens and the prevention of serious health outcomes from GI infection for some groups but also raises further questions about protection across all members of the community/ population. This alongside previous evidence highlights the need for revision of resource allocation, in addition to a place-based approach which takes into account, the relative need, resource deficits and vulnerabilities on areas.

### Conclusion

This study was conducted at the at the end of a period of sustained cuts to local authority budgets that hit the most deprived areas hardest. Overall, we show that the relationship between funding and GI outcomes is complex and multifaceted, with no one clear directionally relationship between expenditure and GI infections. We do show increased resource allocation related to expenditure for food safety services may help prevent illness from foodborne pathogens for some groups, and severe GI infection illness for high exposure risk age groups. However, more work is needed to understand the nuanced patterns by age and why the opposite trend is observed for other age groups.

Simultaneously, those from poorer communities are experiencing higher rates of GI infection and worse outcomes; in addition to our findings, this underscores both the need for greater understanding of these patterns, and the continued investment in local authority services.

## Data Availability

Local authority administrative data are publicly available. Expenditure data are available from the Place-Based Longitudinal Data Resource (PLDR): https://pldr.org/dataset/cultural-environmental-regulatory-and-planning-services-individu-2omjn
. Data on local authority food law enforcement activity, including staffing and establishment numbers, are available from data.gov.uk: https://www.data.gov.uk/dataset/090b5b23-5020-4480-96a0-8b294ca82653/local_authority_food_law_enforcement_returns
. Data on the number of interventions were obtained via Freedom of Information requests from the Food Standards Agency.
Health outcome data used in this study are not publicly available due to data governance and confidentiality restrictions.

https://www.data.gov.uk/dataset/090b5b23-5020-4480-96a0-8b294ca82653/local_authority_food_law_enforcement_returns

## Acknowledgements

We gratefully acknowledge the contributions of Alexandros Alexiou, David Hughes, Roberto Vivancos, Valerie Decraene, Richard Dunn, Nicola Love, Daras, Konstantinos, Simon Melican, Samantha Walters, Jane Muizelaar, Roger Gibb.

## Contributors

LM, HEC, XZ, MAG, MAC, IEB, BB and DH were involved in conceptualisation of the study. LM and DH were involved in investigation, and project administration. LM, DH, HEC, MAG, BB, IB and XZ contributed to methodology. LM carried out visualisation, writing–original draft preparation. LM, HEC, XZ, MAG, MAC, IEB, BB and DH contributed to writing–reviewing and editing. HEC, XZ, MAG, MAC, IEB, BB and DH contributed to supervision. BB was responsible for funding acquisition. LM accepted full responsibility for the finished work and/or the conduct of the study, had access to the data and controlled the decision to publish. LM is responsible for the overall content as guarantor

## Competing interests

DH is currently in receipt of research grant support from the Food Standards Agency. IB was AstraZeneca’s Chief Data Scientist Advisor 2019-2023. LM, RG, MAC, HEC, XZ, MAG, and BB declare no relevant competing interests.

*Ethical approval for this work has been granted by the ethics committee at the university of Liverpool*

