## Supplementary material for "Understanding the impact of local authority resource allocation on gastrointestinal infections in England": Online Supplemental material

### 1 Data background

Table 1 Data background table, variables and sources

| Variable | Source |
| --- | --- |
| FTE(LAEMS) | Open access: <https://www.data.gov.uk/dataset/090b5b23-5020-4480-96a0-8b294ca82653/local-authority-food-law-enforcement-returns> |
| Number of interventions achieved (LAEMS) | FOI request from FSA |
| Food Safety expenditure , FSIC expenditure, ER expenditure | Open access: <https://pldr.org/dataset/2omjn/cultural-environmental-regulatory-and-planning-services-individual-spending-lines-r05-fin0761> |
| Laboratory confirmed pathogen data | Protected data accessed with data sharing agreement : Second Generation Surveillance System |
| Number of hospital admissions due to GI infection | Protected data accessed with data sharing agreement: Hospital Episode Statistics |
| Number of NHS 111 calls for diarrhoea an d vomiting | Protected data accessed with data sharing agreement: NHS 111 |

##### Number of interventions achieved

We look at the annual number of interventions carried out per establishment. Interventions include inspections, sampling, audits, monitoring, surveillance and intelligence gathering, as well as advice and education (33).

##### Food Hygiene full time equivelant (FTE)

###### *1.3 Local authority enforcement monitoring scheme FTE data*

The number of FTE reflect the proportion of time spent by staff on food hygiene issues (1). It is important to note there is no prescriptive guidance on how the time spent on enforcement should be determined, therefore figures supplied are estimates (1). Due to the variety of methods on calculating time the FSA only use the data to compare year on year to look at overall trends in FTE across the UK (1). Here in this paper, we do not make direct comparisons between LA’s, complying with guidance by FSA.

We describe the calculation of the number of FTE. FTE represent the number of professional or administrative staff posts allocated to food law enforcement work and are separated by responsibility e.g. food hygiene or standards (2). LAEMS provide an example of how the FTE are calculated, this is included below:

“*five professional posts were allocated to carry out food hygiene work during the year, and three of these posts were filled for the full twelve months, one for six months and a contractor was employed for 3 months, the figures reported should be as follows: FTE posts allocated = 5; FTE posts occupied = 3.75 i.e. 3 posts filled for full twelve months + one filled for six months (0.5) + one contractor employed for three months (0.25). Where a professional and/or administration staff member only spends a proportion of their time on food hygiene and/or food standards issues, the calculation should reflect this. We recognise that the figures supplied will often be ‘educated estimates’.” (Local authority enforcement monitoring system 2019*) (2)

### 2 Missing data

Table 2 lists the local authorities removed from analysis underneath the respective reasons for removal.

Table 2 Local authorities removed for analysis

|  | **LA’s removed due to 50% or more missing** | **LA’s removed due to reporting with food standards** | **LA’s removed due to inconsistent reporting** |
| --- | --- | --- | --- |
| 1 | Stratford Upon Avon | Westminster | Adur |
| 2 | Rutland | Hillingdon | Worthing |
| 3 | Rochdale | Bexley | Rother |
| 4 | Medway | Hartlepool | Wealden |
| 5 | Tameside | Bolton | Babergh |
| 6 | High peak |  | Midsuffolk |
| 7 | Herefordshire |  |  |
| 8 | Hambleton |  |  |
| 9 | Craven |  |  |
| 10 | Portsmouth |  |  |
| 11 | Epsom and Ewell |  |  |
| 12 | Hinckley Bosworth |  |  |
| 13 | Hart |  |  |

*Note: The following local authorities provided no service expenditure data for the specified years; Havant 2017, East Hampshire 2017, Newark and Sherwood 2012, 2013 and Folkestone Hythe for 2017 and 2018.*

*Adur, Worthing, Babergh. Midsuffolk, Rother and Wealden were also excluded due to inconstant reporting level*

It was identified that negative expenditure was present in a small number of local authorities. We treated these data as 0/missing after contacting a sample of the authorities and identifying mixed reasons as to why there may be negative values reported including error, and system calculations and general uncertainty. Therefore, we felt the best course of action would be to treat the data as missing for multiple imputation models.

### S3 Sensitivity analysis

#### S3.1 No missing data results

| **Exposure** | **Lab confirmed Campylobacter** | **Lab confirmed Salmonella** | **Lab confirmed E. Coli O157** |
| --- | --- | --- | --- |
| **- £1 food safety** | 1.003 0.983 1.024  P = 0.7627 | 1.008 0.981 1.035  P = 0.5698 | 1.033 0.980 1.088  P = 0.2244 |
| **£1 FSIC expenditure** | 0.997 0.991 1.004  P = 0.4671 | 1.005 0.995 1.015  P = 0.3719 | 0.990 0.972 1.009  P = 0.3156 |
| **FTE per 10,000 of the population?** | 1.190 0.933 1.518  P = 0.1613 | 1.311 0.964 1.784  P = 0.0847 | 0.995 0.487 2.034  P = 0.9899 |

| **Exposure** | **Admission age all age**  **IRR 95% CI** | **Admission age 5-9**  **IRR 95% CI** | **Admissions age 10 – 19**  **IRR**  **95% CI 10 – 19** | **Admission age 20-59 IRR**  **95% CI** | **Admission age 60-74 IRR**  **95% CI** | **Admission age 75 +  IRR**  **95% CI** |
| --- | --- | --- | --- | --- | --- | --- |
| **£1 Food safety** | 1.003 (0.992, 1.013) p = 0.6245 | 1.028 (0.998, 1.059)  p= 0.0635 | 1.022 (1.004, 1.040) p = 0.0183 | 0.984 (0.972, 0.997)p = 0.0160 | 0.983 0.964 1.002  P = 0.0803 | 1.006 (0.983, 1.029)  P= 0.6185 |
| **£1 FSIC expenditure** | 1.002 (0.998, 1.006)  P= 0.2322 | 1.005 0.994 1.01  P= 0.3892 | 1.003 (0.996, 1.010)  P= 0.4398 | 0.996 0.991 1.001  P= 0.0833 | 1.000 0.992 1.008  P= 0.9858 | 1.009 1.001 1.017  P= 0.0306 |
| **FTE per 10,000 of the population** | 1.069 (0.925, 1.235)P= 0.3688 | 2.394 1.636 3.504  P= 0.0000 | 1.521 1.151 2.010  0.0032 | 0.879 0.741 1.044  0.1427 | 0.657 0.490 0.88  P= 0.0050 | 0.825 0.593 1.149  P= 0.2553 |

| **Exposure** | **NHS 111 Calls**  **IRR all**  **95% CI** | **NHS 111 Calls age 5-9**  **IRR 95% CI** | **NHS 111 Calls age 10 -19**  **IRR 95% CI** | **NHS 111 Calls age 20-59 IRR**  **95% CI** | **NHS 111 Calls age 60-74 IRR**  **95% CI** | **NHS 111 Calls age 75 +  IRR**  **95% CI** |
| --- | --- | --- | --- | --- | --- | --- |
| **£1 Food safety** | 0.990 (0.964, 1.01)  P= 0.4915 | 0.973 (0.944, 1.002)  P= 0.0656 | 0.994 (0.966, 1.022)  P= 0.6533 | 1.001 (0.972, 1.030)  P= 0.9729 | 0.997 (0.969, 1.026)   p= 0.8470 | 0.991 (0.959, 1.025)   p= 0.6098 |
| **£1 FSIC expenditure** | 0.998 (0.979, 1.018)  P= 0.8665 | 0.985 (0.965, 1.006)  P=0.1573 | 0.995 (0.976, 1.014)  P= 0.5972 | 0.998 (0.978, 1.018)  P= 0.8353 | 0.997 (0.975, 1.020)  P= 0.7852 | 0.999 (0.973, 1.026)  P=0.9340 |
| **FTE per 10,000 of the population** | 0.643 (0.356, 1.163)  P= 0.1444 | 0.542 (0.285, 1.030)  P= 0.0616 | 0.668 (0.356, 1.252)  P= 0.2081 | 0.638 (0.345, 1.182)  P= 0.1533 | 0.676 (0.370, 1.237)  P= 0.2042 | 0.704 (0.361, 1.373)  P= 0.3031 |

#### S3.2 NHS 111 June-Dec

| **Exposure** | **NHS 111 Calls**  **IRR all**  **95% CI** | **NHS 111 Calls age 5-9**  **IRR 95% CI** | **NHS 111 Calls age 10 -19**    **IRR 95% CI** | **NHS 111 Calls age 20-59 IRR**  **95% CI** | **NHS 111 Calls age 60-74 IRR**  **95% CI** | **NHS 111 Calls age 75 +  IRR**  **95% CI** |
| --- | --- | --- | --- | --- | --- | --- |
| **£1 Food safety** | 0.9904 0.9666 1.0147 p= 0.4349 | 0.9772 0.9512 1.0039  0.0938 | 0.9939 0.9688 1.0197  P= 0.6396 | 0.9995 0.9735 1.0261  P= 0.9692 | 0.9943 0.9696 1.0196  p= 0.6543 | 0.9861 0.9562 1.0170  p= 0.3735 |
| **£1 FSIC expenditure** | 0.9972 0.9797 1.0150  P= 0.7524 | 0.9876 0.9696 1.0059  p= 0.1838 | 0.9965 0.9798 1.0136  P=0.6889 | 0.9993 0.9823 1.0166 p= 0.9333 | 0.9989 0.9792 1.0190 p= 0.9145 | 0.9996 0.9760 1.0238  P= 0.9726 |
| **FTE per 10,000 of the population** | 0.6461 0.3577 1.1671   p= 0.1477 0 | 0.5422 0.2854 1.0300 p= 0.0615 | 0.6717 0.3587 1.2579 p= 0.2138 | 0.6405 0.3462 1.1850 p= 0.1558 | 0.6785 0.3714 1.2395  p= 0.2071 | 0.7079 0.3633 1.3791  P= 0.3099 |

#### S3.3 Analysis by deprivation, most deprived compared to least deprived

| **Exposure** | **Admission Q1** | **Admission Q5** |
| --- | --- | --- |
| **£1 Food safety** | 1.0019 0.9852 1.0190 p = 0.8224 | 1.0214 0.9966 1.0467 p= 0.0909 |
| **£ FSIC expenditure** | 0.9986 (0.9906,1.0067) p= 0.7322 | 0.9995 0.9923 1.0067 p= 0.8920 |
| **FTE per 10,000 of the population** | 1.1086 (0.8637,1.4229)  P= 0.4181 | 1.2609 (0.8879 1.7905)  p= 0.1952 |
| **Exposure** | **NHS111 Q1** | **NHS 111 Q5** |
| **£1 Food safety** | 1.0509 0.9253 1.1935  P= 0.4445 | 0.9914 0.9397 1.0459  P= 0.7507 |
| **£ FSIC expenditure** | 1.0468 0.9580 1.1439  P= 0.3118 | 0.9816 0.9391 1.0261  P= 0.4117 |
| **FTE per 10,000 of the population** | 0.5805 0.2739 1.2302 p= 0.1558 | 2.2099 0.7733 6.3152 p= 0.1388 |

| **Exposure** | **Lab confirmed *Campylobacter* per 100,000**  **IRR**  **95% CI Q1** | **Lab confirmed *Campylobacter* per 100,000**  **IRR**  **95% CI Q5** |
| --- | --- | --- |
| **£1 Food safety** | 0.9960 (0.9703, 1.0223) p = 0.7607 | 0.96 (0.94, 0.99) p = 0.0052 |
| **£ FSIC expenditure** | 0.9974 (0.9823, 1.0128) P = 0.7406 | 1.0042 0.9947 1.0138 p = 0.3863 |
| **FTE per 10,000 of the population** | *1.972 (0.7398, 1.9376)*  *P = 0.4636* | 0.9810 (0.4548, 2.1159) p = 0.9609 |
| **Exposure** | **Lab confirmed *Salmonella* per 100,000**  **IRR**  **95% CI Q1** | **Lab confirmed *Salmonella* per 100,000**  **IRR**  **95% CI** |
| **£1 Food safety** | 0.95 (0.91, 0.99 ) p = 0.0181 | *0.9940 (0.9554, 1.0342) p =0.7666* |
| **£ FSIC expenditure** | 0.9949 0.9699 1.0206 p = 0.6967 | 1.0017 0.9893 1.0141 P = 0.7933 |
| **FTE per 10,000 of the population** | *0.7398 (0.3437, 1.5922) p =0.4409* | 0.6143 (0.2785, 1.3551) p =0.2274 |
| **Exposure** | **Lab confirmed *E.coli* per 100,000**  **IRR**  **95% CI Q1** | **Lab confirmed *E.coli* per 100,000**  **IRR**  **95% CI** |
| **£1 Food safety** | 0.9278 (0.8586, 1.0026) P = 0.0582 | 0.9438 (0.7295, 1.220)  P = 0.6595 |
| **£ FSIC expenditure** | 0.968 (0.937, 0.999) p = 0.0426 | 0.9691 (0.9185 1.0224) P = 0.2502 |
| **FTE per 10,000 of the population** | 0.092 (0.0171, 0.49)  P = 0.0053 | 2.2409 (0.4295, 11.6917)  P = 0.3384 |

### S3.4 Negative binomial model results for Interventions and ER per capita

We exclude analysis for ER expenditure and the number of interventions from our main reporting. Total ER expenditure, while capturing important activities related to public health, also encompasses services unrelated to infection control or GI illness. Any conclusions therefore reflect unrelated spending lines. The number of interventions per establishment is an indicator of service delivery, how often an establishment receives an intervention is determined by risk faced to the public (47). Therefore, increased interventions may reflect increased need over increased capacity and consequently may be an example of reverse causality.

#### Results

The number of interventions per establishment had no significant relationship with the rate of laboratory – confirmed pathogens. Environmental and regulatory expenditure was associated with a significant reduction in the rate of E. coli O157 infections (IRR = 0.996, 95%CI: 0.993, 0.9990).

| **Exposure** | **Lab confirmed Campylobacter per 100,000**  **IRR**  **95% CI** | **Lab confirmed Salmonella   100,000**  **IRR 95% CI** | **Lab confirmed E. coli   100,000**  **IRR 95% CI** |
| --- | --- | --- | --- |
| **Interventions per establishment** | 1.0238 (0.9368 1.1187) p = 0.6040 | 0.9700 (0.8748 1.0755) p = 0.5627 | 1.0048 (0.7833, 1.2890) p = 0.9697 |
| **£ ER expenditure** | 1.0002 (0.9988 1.0015)  p=0.7894 | 1.0009 (0.9991 1.0027) p = 0.3218 | 0.996(0.993, 0.9990) *p = 0.009*   1.00(0.99, 1) *p = 0.009* |

There was no significant relationship between the number of interventions per establishment and the number of hospital admissions. One unit increase in ER was associated with an increase in hospital admissions of the population (IRR = 1.0018; 95% CI: 1.0007,1.0029

| **Exposure** | **Admission per 100,000 allage**  **IRR 95% CI** | **Admission per 100,000 age 5-9**  **IRR 95% CI** | **Admissions per 100,000**  **IRR**  **95% CI 10 – 19** | **Admission per 100,000 age 20-59 IRR**  **95% CI** | **Admission per 100,000 age 60-74 IRR**  **95% CI** | **Admission per 100,000 age 75 +  IRR**  **95% CI** |
| --- | --- | --- | --- | --- | --- | --- |
| **Interventions per establishment** | 1.0290 0.9599 1.1030  P= 0.4207 | 1.1145 0.9176 1.3538  P= 0.2744 | 1.0625 0.9137 1.2354  P= 0.4309 | 0.9701 0.8953 1.0512  P= 0.4591 | 1.0179 0.8985 1.1531  P= 0.7805 | 1.0107 0.8958 1.1403  P= 0.8630 |
| **£ ER expenditure** | 1.0004 0.9997 1.0011  0.2685 | 0.9991 0.9978 1.0004  0.1730 | 0.9999 0.9990 1.0007  P= 0.7518 | 0.9997 0.9988 1.0006  P= 0.5733 | 1.0010 0.9999 1.0022  P= 0.0839 | 1.0018 1.0007 1.0029  P= 0.0015 |

There was no significant association between the number of interventions per establishment and the rate of NHS 111 calls, or ER expenditure and the rate of NHS 111 calls.

| **Exposure** | **NHS 111 Calls per 100,000 all age**  **IRR 95% CI** | **NHS 111 Calls per 100,000 age 5-9**  **IRR 95% CI** | **NHS 111 Calls per 100,000**  **IRR 10 -19**  **95% CI** | **NHS 111 Calls per 100,000 age 20-59 IRR**  **95% CI** | **NHS 111 Calls per 100,000 age 60-74 IRR**  **95% CI** | **NHS 111 Calls per 100,000 age 75 +  IRR**  **95% CI** |
| --- | --- | --- | --- | --- | --- | --- |
| **Interventions per establishment** | 1.0910 (0.9543, 1.2474)  P= 0.2024 | 1.0895 (0.9406, 1.2621) p= 0.2530 | 1.0182 (0.8890, 1.1662) p = 0.7945 | 1.0812 (0.9347, 1.2508) p= 0.2933 | 1.1229 (0.9510, 1.3259) p = 0.1715 | 1.1713 (0.9519, 1.4412) p = 0.1351 |
| **£ ER expenditure** | 0.9989 0.9969 1.0009 P = 0.2764 | 0.9980 0.9959 1.0001  P= 0.0613 | 0.9995 0.9974 1.0016  P= 0.6558 | 0.9995 0.9973 1.0016  P= 0.6320 | 0.9991 0.9971 1.0011  P= 0.3995 | 0.9980 0.9959 1.0001  P= 0.0563 |

### S3.5 Analysis using 1 year lag

This method allows us to analyse the previous year's expenditure and staff allocation with current year health outcomes, to understand if the previous year's resource allocation influenced health outcomes of the following year. This analysis identified no significant findings, indicating that overall, the previous year’s resource allocation had no impact on the health outcomes of the following year.

| **Exposure** |  | **Lab confirmed Campylobacter** | **Lab confirmed Salmonella** | **Lab confirmed E. Coli O157** |
| --- | --- | --- | --- | --- |
| **- £1 food safety** |  | 1.0035 0.9844 1.0231 p= 0.7194 | 1.0019 0.9773 1.0271  P= 0.8833 | 1.0471 0.9990 1.0975 p= 0.0549 |
| **£1 FSIC expenditure** |  | 1.0010 0.9939 1.0081 p= 0.7830 | 1.0028 0.9932 1.0124  P= 0.5711 | 0.9828 0.9665 0.9994  P= 0.0421 |
| **FTE per 10,000 of the population?** |  | 1.0418 0.8212 1.3218  P= 0.7357 | 1.2787 0.9801 1.6682  0.0700 | 1.5092 0.7569 3.0095 p= 0.2425 |

| **Exposure** | **Admission age all age**  **IRR 95% CI** |
| --- | --- |
| **£1 Food safety** | 1.0044 0.9947 1.0142  P= 0.3789 |
| **£1 FSIC expenditure** | 1.0026 0.9990 1.0063  P= 0.1591 |
| **FTE per 10,000 of the population** | 1.0489 0.9172 1.1996  P= 0.4854 |

| **Exposure** | **NHS 111 Calls**  **IRR all**  **95% CI** |
| --- | --- |
| **£1 Food safety** | 0.9904 0.9679 1.0135 P= 0.4118 |
| **£1 FSIC expenditure** | 0.9995 0.9835 1.0157 p= 0.9497 |
| **FTE per 10,000 of the population** | 0.6681 0.4075 1.0952 p= 0.1097 |
